# Pre-Injury Frailty and Clinical Care Trajectory of Geriatric Trauma Patients: A Retrospective Cohort Analysis of A Large Level I US Trauma Center

**DOI:** 10.1101/2023.06.19.23291575

**Authors:** Oluwaseun Adeyemi, Corita Grudzen, Charles DiMaggio, Ian Wittman, Ana Velez-Rosborough, Mauricio Arcila-Mesa, Allison Cuthel, Helen Poracky, Polina Meyman, Joshua Chodosh

**Author notes:** Corresponding Author* Dr. Oluwaseun John Adeyemi, Ronald O Perelman Department of Emergency Medicine, New York University Grossman School of Medicine, New York, USA. Funding: This study was not funded. Disclosures: The authors declare no competing interests.

## Abstract

**Background:** Pre-injury frailty among older trauma patients is a predictor of increased morbidity and mortality. We sought to determine the relationship between frailty status and the care trajectories of older adult patients who underwent frailty screening in the emergency department (ED).

**Methods:** Using a retrospective cohort design of a single institutional trauma database, we pooled data on trauma patients, 65 years and older, who had frailty screening at ED presentation (N=987). The predictor variable was frailty status, measured as either robust, pre-frail, or frail. The outcome variables were measures of clinical care trajectory: inpatient admission, length of hospital stay, home discharge, and discharge to rehabilitation. We controlled for age, sex, race/ethnicity, body mass index, Charlson Comorbidity Index, injury type and severity, and Glasgow Coma Scale score. We performed multivariable logistic and quantile regressions to measure the influence of frailty on post-trauma care trajectories.

**Results:** The mean (SD) age of the study population was 81 (9.0) years and the population was predominantly female (66%) and non-Hispanic White (64%). Compared to older adult trauma patients classified as robust, those categorized as frail had 2.8 (95% CI: 1.75 – 4.40), 0.4 (95% CI: 0.27 – 0.63), and 2.1 (95% CI: 1.38 – 3.27) times the adjusted odds of hospital admission, home discharge, and discharge to rehabilitation, respectively. Those classified as pre-frail (Adjusted MD: 1.0; 95% CI: 0.46 – 1.54) and frail (Adjusted MD: 2.0; 95% CI: 1.35 – 2.65) had longer lengths of hospital stay compared to those classified as robust.

**Conclusion:** Pre-injury frailty is a predictor of care trajectories for older-adult trauma patients.

## Introduction

Frailty is a clinical syndrome comprising weakness, slowness, diminished physical activities, exhaustion, and weight loss.^1, 2^ It is a chronically acquired clinical state that manifests with increased vulnerability to dependency and disability when exposed to physiological and external stressors.^3, 4^ It is estimated that 10 to 15% of community-dwelling older adults are frail,^5, 6^ and among older adult trauma patients, preinjury frailty prevalence ranges from two to 33%.^7^ Frailty not only predisposes older adults to injuries such as falls^8, 9^ but also increases injury-associated morbidity and mortality.^7^ While the natural history of frailty includes the potential of reversal and improvement,^4^ frailty tends to progress more during acute stress conditions such as traumatic injuries.^10^ The inability to mount an adequate physiologic response to trauma leads to worsening weakness, weight loss, and diminished physical activities which culminate in the loss of one or more domains of activities of daily living and disability.^5, 11^

Earlier studies have reported the association between pre-injury frailty and morbidity and mortality among older adults.^12-14^ However, little is known about the role pre-injury frailty plays in the clinical care trajectory among older adult trauma patients. Earlier studies have reported that hospitalized older adults spend more than 80 percent of their hospital stay lying in bed,^15, 16^ and approximately 20 percent lose their ability to walk unassisted at discharge.^17^ Sarcopenia, a pathologic feature of physical frailty and clinically manifested as loss of muscle mass,^18^ develops as early as within the first 72 hours of patient admission.^15^ It is, therefore, possible that without an intent to manage frailty as a comorbid illness, pre-injury frailty and/or frailty progression may influence the clinical care trajectory of older adult trauma patients.

While frailty cannot be corrected during a single inpatient admission, it can be managed. Identifying and managing pre-injury frailty among older adult trauma patients can aid in slowing down clinical frailty progression through early initiation of nutritional rehabilitation, exercise physiotherapy, and early mobilization.^10, 19^ We hypothesized that, in the absence of deliberate interventions to manage frailty during an index hospital stay, older adult trauma patients with pre-injury frailty who present at the emergency department (ED) will be more likely to be admitted to inpatient units, will have longer hospital stays, be less likely to be discharged home and more likely to be discharged to rehabilitation centers such as acute or subacute rehabilitation centers or skilled nursing facilities. This study, therefore, aims to assess the association between pre-injury frailty and the care trajectory experienced by older adult trauma patients during an index ED visit.

## Methods

### Study Design and Population

For this retrospective cohort study, we pooled trauma data from the institutional trauma registry of a large urban level I trauma center that serves a racially and ethnically diverse population. The study population was older adult trauma patients who presented to the ED between August 2020 and December 2021.

### Inclusion and Exclusion Criteria

Across the selected years, a total of 1,676 older adult trauma patients presented to the ED with traumatic injuries (Figure 1).. Also, a total of 645 (38% of 1,676) patients, which included four patients who died in the ED, did not have a documented frailty screening. We excluded this unscreened population. A total of 34 patients had more than one ED visit during the study period. All recurrent ED visits (n= 37) were fall-related. We selected only the most recent visit (excluding 37; 2% of 1,676). Finally, we performed a listwise deletion of variables with missing values (n=7; <1% of 1,676). The final dataset was 987 (36% of 2,823) older adults with trauma injuries who were screened for frailty.

**Figure 1:**
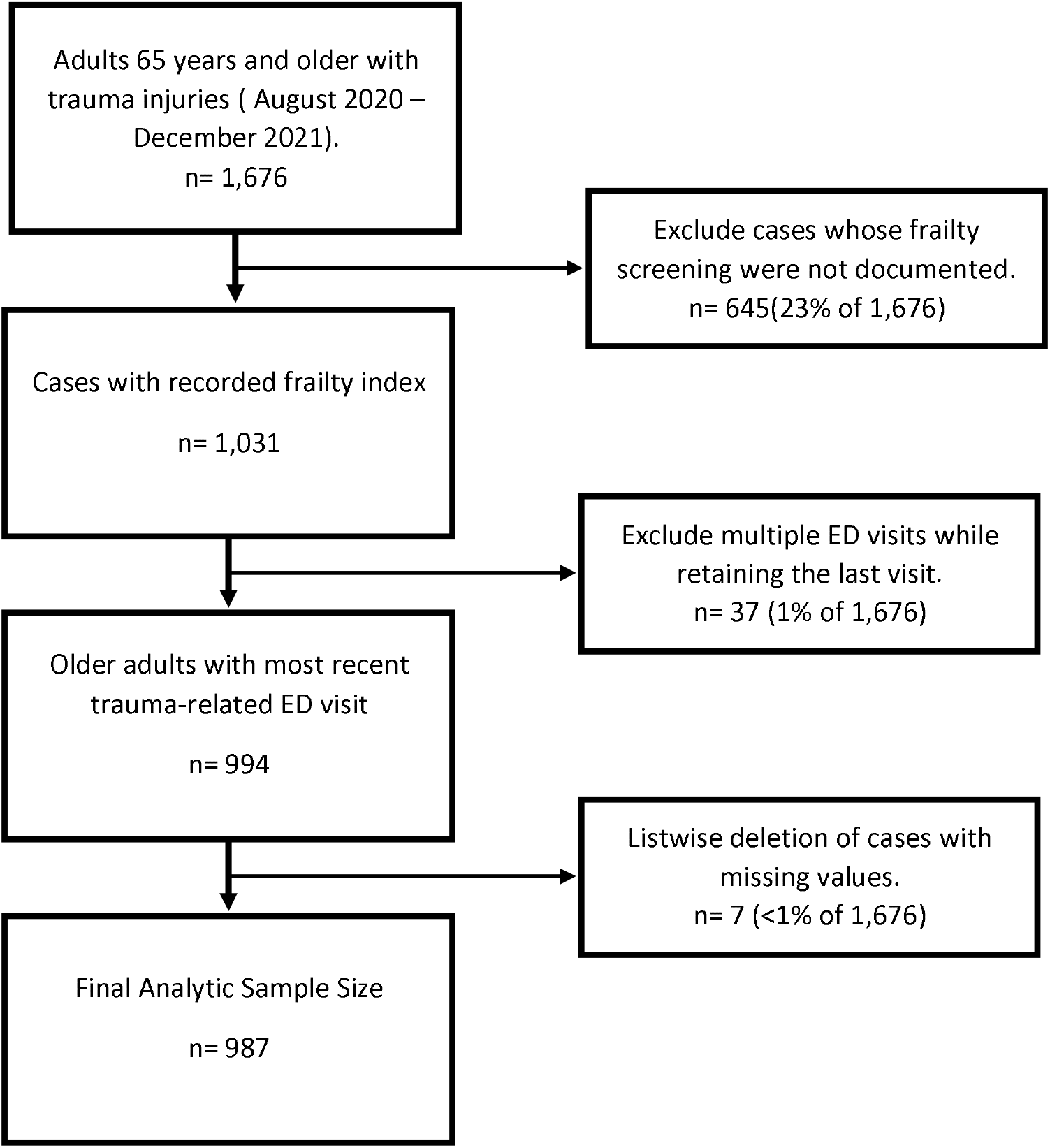
Data selection steps.

### Outcome Variables

Clinical care trajectory variables were the outcomes of interest. We measured clinical care trajectories using four variables: (1) inpatient admission, (2) length of hospital stay, (3) home discharge, and (4) discharge to rehabilitation. We defined inpatient admission as an ED disposition that required transfer from the ED to an inpatient unit. Length of hospital stay was defined as the duration (in days) from inpatient admission to time of discharge order from the index hospital or transfer order to another level I hospital. Patients who were discharged from the ED were assigned a value of zero for the in-hospital length of stay. We defined home discharge as a hospital disposition that allows the patient to go home with or without home health services. We defined discharge to rehabilitation as any hospital disposition that requires the transfer of the patient to an acute or subacute inpatient rehabilitation or a new placement in a skilled nursing facility. All four measures, except the length of hospital stay, were measured as binary variables. The length of hospital stay was measured as a continuous variable.

### Predictor Variable

The predictor variable was frailty status, assessed at ED presentation. Frailty status was defined using the FRAIL index - an acronym for Fatigue, Resistance, Ambulation, Illnesses, and Loss of weight.^3, 20^ The FRAIL index has good face and construct validity and reliability of 0.53.^21^ Each of the five items in the FRAIL index is measured as a binary variable (yes = 1, no = 0). The FRAIL score, therefore, ranges from 0 to 5. Consistent with the FRAIL index scoring, we generated three ordered categories from the scores: Robust (a score of 0), Pre-frail (scores 1 to 2), and Frail (scores 3 to 5) (Appendix 1).^22, 23^

### Potential Confounders

We controlled for age, sex, race/ethnicity, body mass index, injury mechanism, recurrent fall injury, injury severity, Charlson Comorbidity Index, and the Glasgow Coma Scale at the ED presentation. Age was measured as a continuous variable while sex was measured as a binary variable. Race/ethnicity was measured as a four-level categorical variable of non-Hispanic White, non-Hispanic Black, Hispanic, and other races. Body mass index was measured as an ordered variable of underweight (<18.5 kg/m^2^), normal weight (18.5 – 24.9 kg/m^2^), overweight (25 – 29.9 kg/m^2^), and obese (≥ 30.0 kg/m^2^). We defined injury mechanism as either fall or non-fall related. Recurrent fall injury was measured as a binary variable and defined as the occurrence of ED presentation on account of more than one fall-related injury during the study period. We defined the injury severity using the Injury Severity Scale (ISS) score. The ISS score, computed using the abbreviated injury score of the top three injured body regions, typically ranges from 0 to 75, with 0 representing no injury and 75 representing non-survivable injury.^24^ Scores of 1-8 represent minor injuries, while scores ranging from 9-15, 16 to 24, and 25 or higher represent moderate, severe, and very severe injuries, respectively.^24^ We defined injury severity as a binary variable – minor and moderate to severe. Also, we defined the Charlson Comorbidity index as a four-level ordered variable of none, one, two, and three or more. The Glasgow Coma Scale score ranges from 3 to 15 and we measured it as a three-level ordered category of mild (13 to 15), moderate (9 to 12), and severe head injury (3 to 8).

### Analysis

We report summary statistics (mean, standard deviation (SD), median, first and third quartile) and frequency distribution of the selected variables and assessed the distribution of variables across the spectrum of robust, pre-frail, and frail categories. Differences across the frailty spectrum were assessed using the Chi-square test, one-way ANOVA, and Kruskal-Wallis test as appropriate. We performed univariable and multivariable logistic regression to assess the association of the frailty categories and reported the odds ratio (OR) and 95% confidence intervals (CI). Also, we performed univariable and multivariable quantile regression to assess the association between the frailty categories and report the median difference (MD) and 95% CI. Lastly, we computed the predicted probabilities of inpatient admission, home discharge, discharge to rehabilitation, and the predicted estimates of the lengths of hospital stay. Data were analyzed using STATA version 17.^25^

## Results

The mean (SD) of the sample population was 81 (9.0) years (Table 1). The population was predominantly female (66%), and non-Hispanic White (65%). Approximately half of the population was either overweight (32%) or obese (18%). Falls accounted for 94% of the injuries, 3% had recurrent fall injuries, and 41% of the population had no co-morbid condition. One third had moderate to severe injuries and 98% had mild head injuries. Sixty-four percent had inpatient admission from the ED and the median (Q1, Q3) length of stay was 2 days (0.0, 5.0). Also, 63% were discharged home and 33% were discharged to rehabilitation.

**Table 1:**
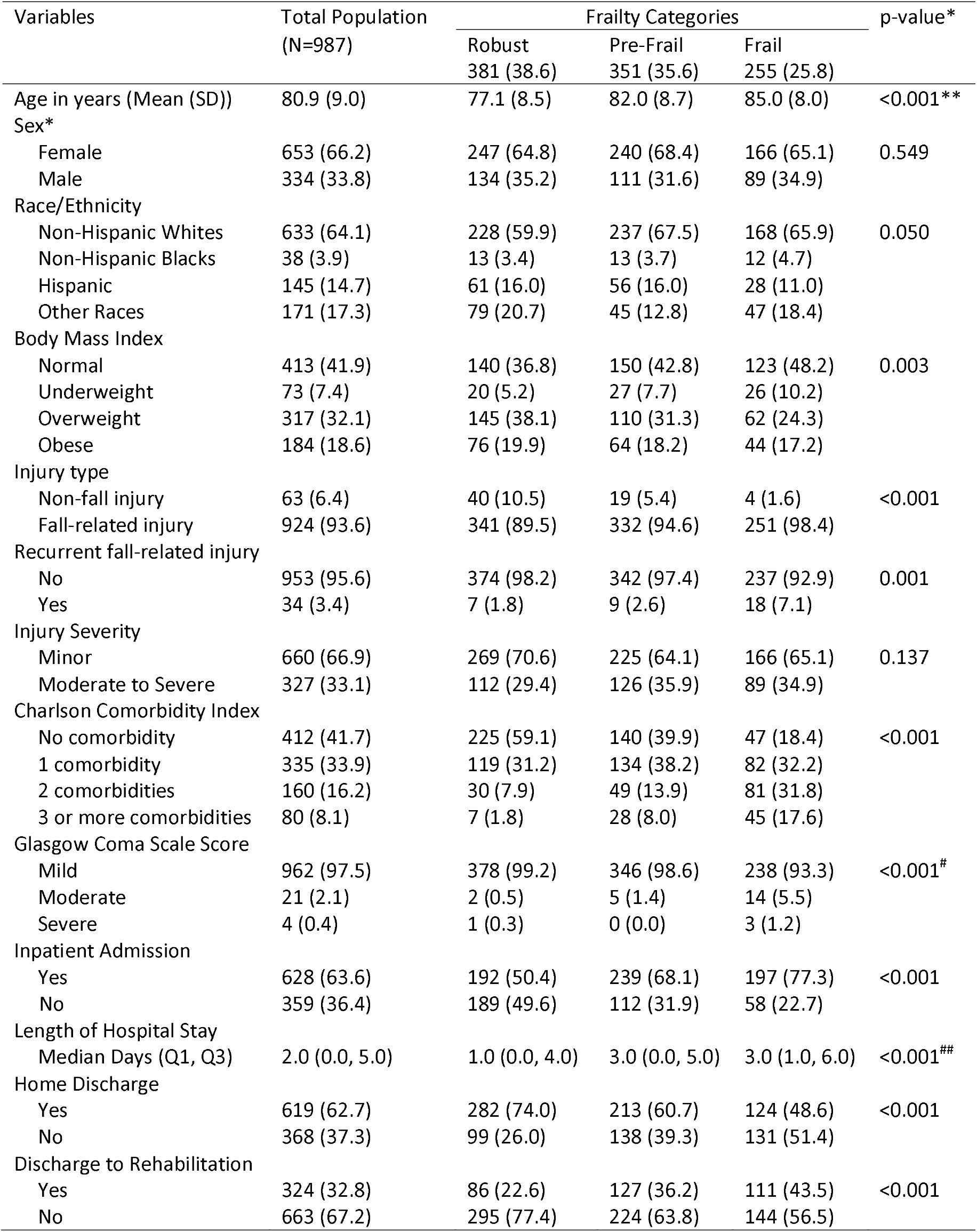
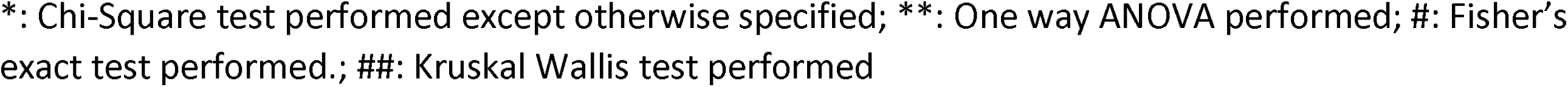
Summary and frequency distribution of the demographic, injury, and measures of care trajectory of older adult trauma patients stratified by frailty category (N=987)

| Variables | Total Population<br>(N=987) | Frailty Categories |  |  | p-value* |
| --- | --- | --- | --- | --- | --- |
|  |  | Robust<br>381 (38.6) | Pre-Frail<br>351 (35.6) | Frail<br>255 (25.8) |  |
| Age in years (Mean (SD)) | 80.9 (9.0) | 77.1 (8.5) | 82.0 (8.7) | 85.0 (8.0) | <0.001** |
| Sex* |  |  |  |  |  |
| Female | 653 (66.2) | 247 (64.8) | 240 (68.4) | 166 (65.1) | 0.549 |
| Male | 334 (33.8) | 134 (35.2) | 111 (31.6) | 89 (34.9) |  |
| Race/Ethnicity |  |  |  |  |  |
| Non-Hispanic Whites | 633 (64.1) | 228 (59.9) | 237 (67.5) | 168 (65.9) | 0.050 |
| Non-Hispanic Blacks | 38 (3.9) | 13 (3.4) | 13 (3.7) | 12 (4.7) |  |
| Hispanic | 145 (14.7) | 61 (16.0) | 56 (16.0) | 28 (11.0) |  |
| Other Races | 171 (17.3) | 79 (20.7) | 45 (12.8) | 47 (18.4) |  |
| Body Mass Index |  |  |  |  |  |
| Normal | 413 (41.9) | 140 (36.8) | 150 (42.8) | 123 (48.2) | 0.003 |
| Underweight | 73 (7.4) | 20 (5.2) | 27 (7.7) | 26 (10.2) |  |
| Overweight | 317 (32.1) | 145 (38.1) | 110 (31.3) | 62 (24.3) |  |
| Obese | 184 (18.6) | 76 (19.9) | 64 (18.2) | 44 (17.2) |  |
| Injury type |  |  |  |  |  |
| Non-fall injury | 63 (6.4) | 40 (10.5) | 19 (5.4) | 4 (1.6) | <0.001 |
| Fall-related injury | 924 (93.6) | 341 (89.5) | 332 (94.6) | 251 (98.4) |  |
| Recurrent fall-related injury |  |  |  |  |  |
| No | 953 (95.6) | 374 (98.2) | 342 (97.4) | 237 (92.9) | 0.001 |
| Yes | 34 (3.4) | 7 (1.8) | 9 (2.6) | 18 (7.1) |  |
| Injury Severity |  |  |  |  |  |
| Minor | 660 (66.9) | 269 (70.6) | 225 (64.1) | 166 (65.1) | 0.137 |
| Moderate to Severe | 327 (33.1) | 112 (29.4) | 126 (35.9) | 89 (34.9) |  |
| Charlson Comorbidity Index |  |  |  |  |  |
| No comorbidity | 412 (41.7) | 225 (59.1) | 140 (39.9) | 47 (18.4) | <0.001 |
| 1 comorbidity | 335 (33.9) | 119 (31.2) | 134 (38.2) | 82 (32.2) |  |
| 2 comorbidities | 160 (16.2) | 30 (7.9) | 49 (13.9) | 81 (31.8) |  |
| 3 or more comorbidities | 80 (8.1) | 7 (1.8) | 28 (8.0) | 45 (17.6) |  |
| Glasgow Coma Scale Score |  |  |  |  |  |
| Mild | 962 (97.5) | 378 (99.2) | 346 (98.6) | 238 (93.3) | <0.001<sup>#</sup> |
| Moderate | 21 (2.1) | 2 (0.5) | 5 (1.4) | 14 (5.5) |  |
| Severe | 4 (0.4) | 1 (0.3) | 0 (0.0) | 3 (1.2) |  |
| Inpatient Admission |  |  |  |  |  |
| Yes | 628 (63.6) | 192 (50.4) | 239 (68.1) | 197 (77.3) | <0.001 |
| No | 359 (36.4) | 189 (49.6) | 112 (31.9) | 58 (22.7) |  |
| Length of Hospital Stay |  |  |  |  |  |
| Median Days (Q1, Q3) | 2.0 (0.0, 5.0) | 1.0 (0.0, 4.0) | 3.0 (0.0, 5.0) | 3.0 (1.0, 6.0) | <0.001<sup>##</sup> |
| Home Discharge |  |  |  |  |  |
| Yes | 619 (62.7) | 282 (74.0) | 213 (60.7) | 124 (48.6) | <0.001 |
| No | 368 (37.3) | 99 (26.0) | 138 (39.3) | 131 (51.4) |  |
| Discharge to Rehabilitation |  |  |  |  |  |
| Yes | 324 (32.8) | 86 (22.6) | 127 (36.2) | 111 (43.5) | <0.001 |
| No | 663 (67.2) | 295 (77.4) | 224 (63.8) | 144 (56.5) |  |

\*: Chi-Square test performed except otherwise specified; \*\*: One way ANOVA performed; #: Fisher's exact test performed.; ##: Kruskal Wallis test performed

Of the 987 patients, 38%, 36%, and 26% were categorized as robust, pre-frail, and frail, respectively. The mean age significantly increased from robust to pre-frail and frail categories (p<0.001) and there were significant differences across the frailty categories by race/ethnicity (p=0.026), body mass index (p=0.001), injury type (p<0.001), recurrent fall injury (p<0.001), Charlson comorbidity index (p<0.001) and Glasgow coma scale score (p<0.001). The proportion of patients that had inpatient admission increased from 51% to 68% and 77% in the robust, pre-frail, and frail categories, respectively. Also, the median (Q1, Q3) lengths of stay increased from 1 (0.0, 4.0) to 2.5 (0.0, 5.0) and 3 (1.0, 6.0) days in the robust, pre-frail, and frail categories, respectively. The proportion of home discharges decreased from 74% to 62% and 48% in the robust, pre-frail, and frail categories, respectively. Additionally, the proportion of discharge to rehabilitation increased from 23% to 36% and 44% in the robust, pre-frail, and frail categories, respectively.

In the unadjusted models, age, fall injuries, having moderate to severe injury, having 2 or more comorbidities and moderate head injury were associated with increased odds of inpatient admission (Table 2). Age, moderate to severe injury, as well as severe head injury were associated with longer hospital stays. Being Hispanic, overweight, or obese was associated with shorter hospital stays. Being a non-Hispanic Black, Hispanic, overweight, or obese was associated with increased odds of home discharge while age, 3 or more comorbidities, and moderate to severe injury were associated with increased odds of discharge to rehabilitation.

**Table 2:**
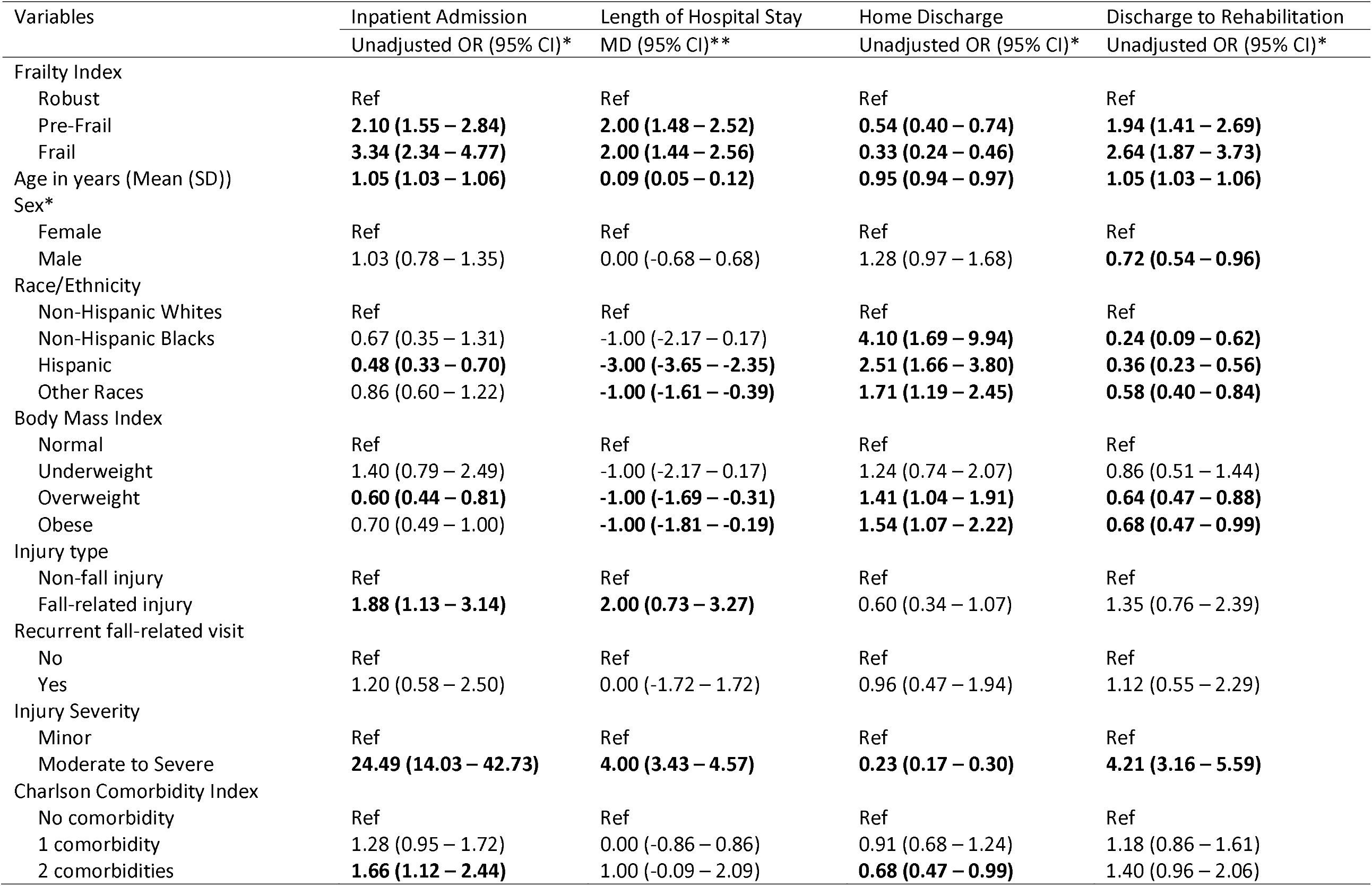
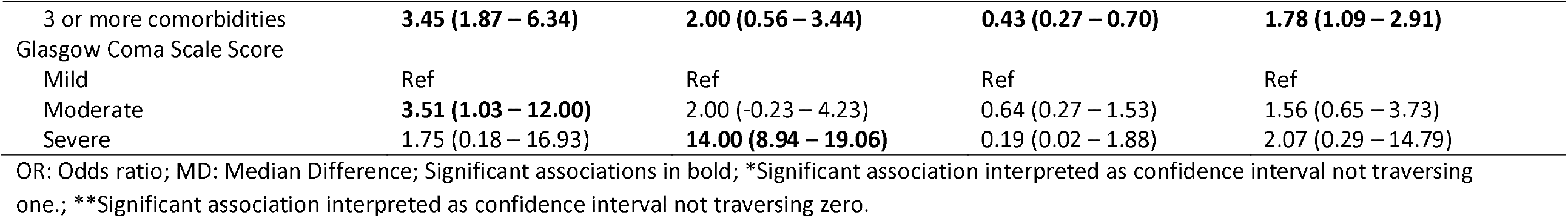
Unadjusted odds of hospital admission, home discharge, discharge to rehabilitation, and median difference in the lengths of hospital stay among older adult trauma patients (N=987)

| Variables | Inpatient Admission<br>Unadjusted OR (95% CI)* | Length of Hospital Stay<br>MD (95% CI)** | Home Discharge<br>Unadjusted OR (95% CI)* | Discharge to Rehabilitation<br>Unadjusted OR (95% CI)* |
| --- | --- | --- | --- | --- |
| Frailty Index |  |  |  |  |
| Robust | Ref | Ref | Ref | Ref |
| Pre-Frail | **2.10 (1.55 – 2.84)** | **2.00 (1.48 – 2.52)** | **0.54 (0.40 – 0.74)** | **1.94 (1.41 – 2.69)** |
| Frail | **3.34 (2.34 – 4.77)** | **2.00 (1.44 – 2.56)** | **0.33 (0.24 – 0.46)** | **2.64 (1.87 – 3.73)** |
| Age in years (Mean (SD)) | **1.05 (1.03 – 1.06)** | **0.09 (0.05 – 0.12)** | **0.95 (0.94 – 0.97)** | **1.05 (1.03 – 1.06)** |
| Sex* |  |  |  |  |
| Female | Ref | Ref | Ref | Ref |
| Male | 1.03 (0.78 – 1.35) | 0.00 (-0.68 – 0.68) | 1.28 (0.97 – 1.68) | **0.72 (0.54 – 0.96)** |
| Race/Ethnicity |  |  |  |  |
| Non-Hispanic Whites | Ref | Ref | Ref | Ref |
| Non-Hispanic Blacks | 0.67 (0.35 – 1.31) | -1.00 (-2.17 – 0.17) | **4.10 (1.69 – 9.94)** | **0.24 (0.09 – 0.62)** |
| Hispanic | **0.48 (0.33 – 0.70)** | **-3.00 (-3.65 – -2.35)** | **2.51 (1.66 – 3.80)** | **0.36 (0.23 – 0.56)** |
| Other Races | 0.86 (0.60 – 1.22) | **-1.00 (-1.61 – -0.39)** | **1.71 (1.19 – 2.45)** | **0.58 (0.40 – 0.84)** |
| Body Mass Index |  |  |  |  |
| Normal | Ref | Ref | Ref | Ref |
| Underweight | 1.40 (0.79 – 2.49) | -1.00 (-2.17 – 0.17) | 1.24 (0.74 – 2.07) | 0.86 (0.51 – 1.44) |
| Overweight | **0.60 (0.44 – 0.81)** | **-1.00 (-1.69 – -0.31)** | **1.41 (1.04 – 1.91)** | **0.64 (0.47 – 0.88)** |
| Obese | 0.70 (0.49 – 1.00) | **-1.00 (-1.81 – -0.19)** | **1.54 (1.07 – 2.22)** | **0.68 (0.47 – 0.99)** |
| Injury type |  |  |  |  |
| Non-fall injury | Ref | Ref | Ref | Ref |
| Fall-related injury | **1.88 (1.13 – 3.14)** | **2.00 (0.73 – 3.27)** | 0.60 (0.34 – 1.07) | 1.35 (0.76 – 2.39) |
| Recurrent fall-related visit |  |  |  |  |
| No | Ref | Ref | Ref | Ref |
| Yes | 1.20 (0.58 – 2.50) | 0.00 (-1.72 – 1.72) | 0.96 (0.47 – 1.94) | 1.12 (0.55 – 2.29) |
| Injury Severity |  |  |  |  |
| Minor | Ref | Ref | Ref | Ref |
| Moderate to Severe | **24.49 (14.03 – 42.73)** | **4.00 (3.43 – 4.57)** | **0.23 (0.17 – 0.30)** | **4.21 (3.16 – 5.59)** |
| Charlson Comorbidity Index |  |  |  |  |
| No comorbidity | Ref | Ref | Ref | Ref |
| 1 comorbidity | 1.28 (0.95 – 1.72) | 0.00 (-0.86 – 0.86) | 0.91 (0.68 – 1.24) | 1.18 (0.86 – 1.61) |
| 2 comorbidities | **1.66 (1.12 – 2.44)** | 1.00 (-0.09 – 2.09) | **0.68 (0.47 – 0.99)** | 1.40 (0.96 – 2.06) |
| 3 or more comorbidities | **3.45 (1.87 – 6.34)** | **2.00 (0.56 – 3.44)** | **0.43 (0.27 – 0.70)** | **1.78 (1.09 – 2.91)** |
| Glasgow Coma Scale Score |  |  |  |  |
| Mild | Ref | Ref | Ref | Ref |
| Moderate | **3.51 (1.03 – 12.00)** | 2.00 (-0.23 – 4.23) | 0.64 (0.27 – 1.53) | 1.56 (0.65 – 3.73) |
| Severe | 1.75 (0.18 – 16.93) | **14.00 (8.94 – 19.06)** | 0.19 (0.02 – 1.88) | 2.07 (0.29 – 14.79) |

OR: Odds ratio; MD: Median Difference; Significant associations in bold; \*Significant association interpreted as confidence interval not traversing one.; \*\*Significant association interpreted as confidence interval not traversing zero.

After adjusting for the potential confounders, pre-injury frailty was significantly associated with the measures of clinical care trajectory (Table 3). Compared to those categorized as robust, patients categorized as pre-frail and frail had 1.9 (95% CI: 1.28 - 2.71) and 2.8 (95% CI: 1.75 - 4.40) times the adjusted odds of inpatient admission. Compared to those categorized as robust, patients categorized as pre-frail and frail had 1.0 (95% CI: 0.46 - 1.54) and 2.0 (95% CI: 1.35 - 2.65) adjusted median increase in their lengths of hospital stays. Compared to those categorized as robust, patients categorized as pre-frail and frail had 34% (AOR: 0.66; 95% CI: 0.47 - 0.95) and 58% (AOR: 0.42; 95% CI: 0.27 - 0.63) reduced odds of home discharge, respectively. Compared to those categorized as robust, patients categorized as pre-frail and frail had 1.6 (95% CI: 1.13 - 2.34) and 2.1 (95% CI: 1.38 - 3.27) times the adjusted odds of discharge to rehabilitation, respectively.

**Table 3:** Adjusted odds and median difference in the measures of care trajectory among older adult trauma patients across the frailty spectrum (N= 987)

| Variables | Inpatient Admission | Length of Hospital Stay | Home Discharge | Discharge to Rehabilitation |
| --- | --- | --- | --- | --- |
|  | Adjusted Odds Ratio (95% CI) | Adjusted Median Difference (95% CI) | Adjusted Odds Ratio (95% CI) | Adjusted Odds Ratio (95% CI) |
| Frailty Index |  |  |  |  |
| Robust | Ref | Ref | Ref | Ref |
| Pre-Frail | **1.86 (1.28 – 2.71)** | **1.00 (0.46 – 1.54)** | **0.66 (0.47 – 0.95)** | **1.62 (1.13 – 2.34)** |
| Frail | **2.78 (1.75 – 4.40)** | **2.00 (1.35 – 2.65)** | **0.42 (0.27 – 0.63)** | **2.13 (1.38 – 3.27)** |

Each model controlled for age, sex, race/ethnicity, body mass index, injury type, recurrent fall injury, injury severity, Charlson comorbidity index, and Glasgow Coma Scale score

The predicted probabilities of inpatient admission increased from 64% (95% CI: 56.9 - 70.4) to 77% (95% CI: 71.2 - 82.0) and 83% (95% CI: 77.6 - 88.3) across the robust, pre-frail, and frail categories (Figure 2). Also, the predicted median lengths of hospital stay increased from 1.3 days (95% CI: 0.9 - 1.6) to 2.3 (95% CI: 1.9 - 2.6) and 3.3 days (95% CI: 2.8 - 3.7) across the robust, pre-frail, and frail categories, respectively. As the predicted probabilities of home discharge reduced from 73% (95% CI: 68.1 - 78.4) to 65% (95% CI: 59.0 - 70.0) and 53% (95% CI: 45.8 - 60.7) across the robust, pre-frail, and frail categories, the predicted probabilities of discharge to rehabilitation increased from 23% (95% CI: 17.7 - 27.3) to 32% (95% CI: 26.8 - 37.3) and 38% (95% CI: 31.0 - 45.3) across the robust, pre-frail, and frail categories.

**Figure 2:**
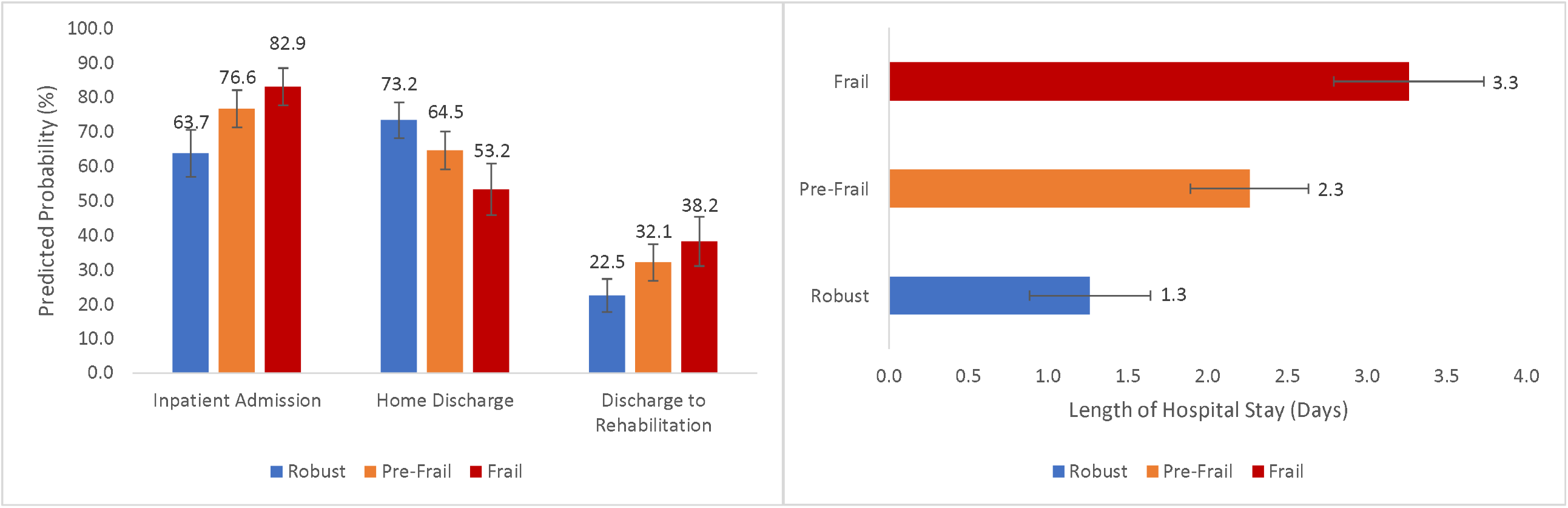
Predicted probabilities of inpatient admission, home discharge, discharge to rehabilitation, and the predicted estimates of the lengths of hospital stay across the robust, pre-frail, and frail spectrum among older adult trauma patients (N=987) Each model controlled for age, sex, race/ethnicity, body mass index, injury mechanism, recurrent fall-related visit, injury severity, Charlson comorbidity index, and Glasgow Coma Scale score

## Discussion

To our knowledge, this is one of the few studies that reports the association between pre-injury frailty and the clinical care trajectory of older adult trauma patients. Earlier studies have reported that frailty increases the odds of inpatient admission, 30-day re-presentation in the ED, and mortality due to an inability to mount up adequate physiologic response to injuries and diseases. ^26-28^ Our study advances this knowledge by demonstrating that frail older adult trauma patients are more likely to be admitted from the ED, stay longer in the hospital, are less likely to achieve home discharge, and are more likely to be discharged to acute rehabilitation centers or skilled nursing facilities. We also report the dose-response pattern in the predicted odds and estimates of these measures of clinical care trajectories across the robust, pre-frail, and frail categories.

Earlier studies have assessed the relationship between frailty and length of hospital stay among the older adult trauma patient population and have provided conflicting results. While studies conducted in developed countries like the United Kingdom, Germany, and Sweden have reported no difference in lengths of hospital stay,^19, 29^ US-based studies have consistently reported longer lengths of hospital stay for those who are frail.^20, 30, 31^ This difference may be a reflection of different health system policies across countries. Studies outside the US report a mean length of hospital stay of 16 to 20 days.^19, 29^ Conversely, the lengths of hospital stay among US older adult trauma patients is three days for non-surgical patients and seven to nine days for those who have surgery.^20, 30, 31^ Additionally, one of Medicare policies, the primary payer of health coverage for older adults, that govern admission into acute rehabilitation facilities is the three-day rule, which requires an inpatient stay of at least three days excluding the day of admission.^32^ Thus, the three-day rule may explain the three-day median stay among frail patients we reported in this study.

Our study shows that older adult trauma patients were less likely to be discharged home but were more likely to be discharged to rehabilitation centers such as acute or subacute care rehabilitation centers or skilled nursing facilities. Discharge to rehabilitation centers has its benefits some of which include access to multidisciplinary care, reduction in unnecessary ED re-presentation and readmission, and access to physical therapy.{Werner, 2019 #16040} Such discharge disposition is, therefore, not a negative disposition but a less preferred option to home discharge. The option of discharging to acute or subacute care rehabilitation centers and skilled nursing facilities will be further deprecated if older adult trauma patients lose one or more domains of activities of daily living during their hospital stay. Disregarding inpatient frailty progression on the assumption that functions lost would be regained or managed in rehabilitation centers is suboptimal care and must be avoided. With skeletal atrophy setting in within 72 hours of immobility,^15, 16^ care plans of older adult trauma patients should include evidence-based interventions such as comprehensive geriatric assessment, exercise and early ambulation, nutritional rehabilitation, and avoidance or limiting the use of tethering devices such as intravenous lines and catheters.^19, 33^

This study has its limitations. Three of the components of the FRAIL index (fatigue, resistance, and ambulation) are self-reported measures. Self-reported bias, therefore, cannot be excluded. Although the FRAIL index has demonstrated strong validity, it has weak reliability. Hence, the lack of consistency when administered by different emergency providers might influence our results. The statistically significant difference we report between the pre-injury frailty and the lengths of hospital stay may lack clinical relevance since the median difference was approximately one day across each category. Our results may, however, have greater relevance to a smaller subset of older adult trauma patients with extended hospital stays (the right-skewed population). Interventions aimed at reducing frailty progression among admitted older adult trauma patients must, therefore, be focused on identifying those likely to have longer stays. Despite these limitations, this study is the first to demonstrate the dose-response patterns in the clinical care trajectory of older adult trauma patients across the frailty spectrum. Future studies should explore the extent to which evidence-based interventions impact frailty progression among admitted older adult trauma patients.

## Conclusion

Pre-injury frailty is associated with increased likelihood of inpatient admission from the ED, prolonged length of hospital stay, reduced discharges to home and increased discharge to rehabilitation among older adult trauma patients. Indeed, it is impossible to correct pre-injury frailty during a trauma admission. However, a lot can be done to slow down or reduce frailty progression through multidisciplinary care shared by the trauma, geriatric, emergency medicine, physiotherapy, social work, case management and nutritional teams. Screening for pre-injury frailty, early and continued inpatient ambulation, and nutritional rehabilitation may slow down frailty progression and improve the quality of care for older adult trauma patients.

## Data Availability

All data produced in the present study are available upon reasonable request to the authors

---

**Appendix 1:**
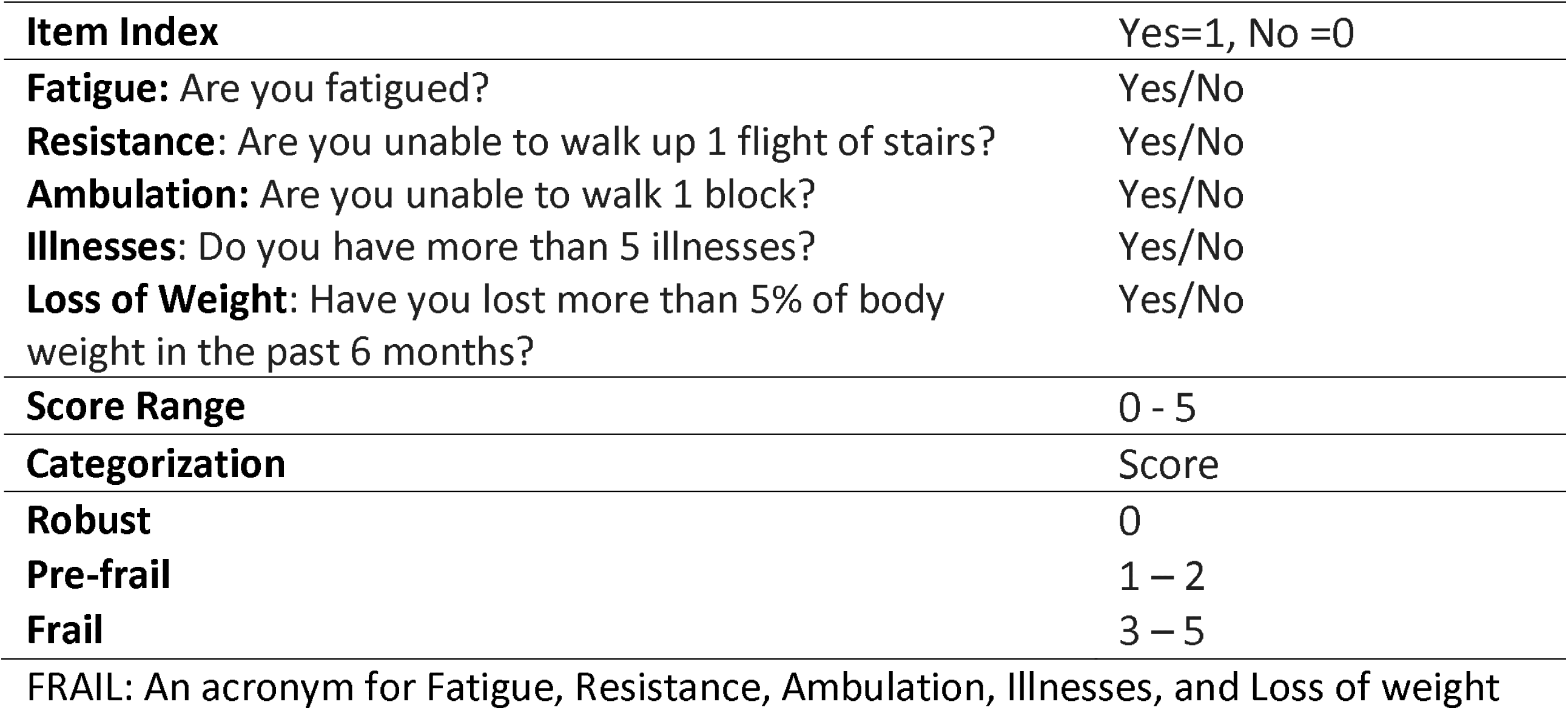
Frailty Index, Categorization, and Scoring.

